# Epigenome-wide analysis identifies methylome profiles linked to obsessive-compulsive disorder, disease severity, and treatment response

**DOI:** 10.1101/2023.02.15.23285944

**Authors:** Rafael Campos-Martin, Katharina Bey, Björn Elsner, Benedikt Reuter, Julia Klawohn, Alexandra Philipsen, Norbert Kathmann, Michael Wagner, Alfredo Ramirez

## Abstract

Obsessive-compulsive disorder (OCD) is a mental disorder affecting 2-3% of the general population. The dynamic nature of epigenetics provides a unique opportunity to find biomarkers of OCD symptoms, clinical progression, and treatment response. Consequently, we analyzed a case-control study on Illumina Methylation EPIC BeadChip from 185 OCD patients and 199 controls. Patients and controls were assessed by trained therapists using the Structured Clinical Interview for DSM-IV. We identified 12 CpGs capable of classifying OCD patients and predicting symptom severity. These CpGs are enriched with **the sweet-compulsive brain hypothesis**, which proposes that OCD patients may have impaired insulin signaling sensitivity due to abnormal dopaminergic transmission in the striatum. Three of the twelve CpG signals were replicated in an independent study reported in the Han Chinese population. Our findings support the role of epigenetic mechanisms in OCD and may help pave the way for biologically-informed and individualized treatment options.

## Introduction

Obsessive-compulsive disorder (OCD) is a psychiatric disorder that affects around 2-3% ^1,2^ of the general population and can result in severe psychosocial impairment if untreated. The disorder is characterized by excessive, unwanted thoughts (obsessions) and/or repetitive behaviors (compulsions) ^3^. Despite OCD’s large burden on affected individuals and the health care system, up to date, no biomarker has been found to classify the disorder in a clinical setting or to aid clinicians to predict response to cognitive-behavioral therapy (CBT).

OCD is considered a multifactorial disorder in which the risk to develop the disease is defined by the complex interaction of genetics, epigenetics, and environmental factors. From a genetic perspective, twin studies have estimated that the heritability of OCD is 47-61% ^4–7^. Despite this high heritability, genome-wide association studies in OCD have identified only one genetic locus reaching genome-wide significance ^8^. This might be explained by the current lack of statistical power to identify genetic variants of small effects. An alternative explanation is that the missing heritability is due to gene x environment interactions contributing to the etiology of OCD ^5,9,10^. For example, research has shown that childhood trauma, stress, or depression, among others, predisposes to OCD, presumably in combination with genetics ^11^. Interestingly, environmental factors are known to exert their effects on disease susceptibility through epigenetic modifications leading to modulation of expression and co-expression of several genes ^12–16^. In humans, the most studied epigenetic modification is the methylation of DNA (DNAm). The development of high throughput array technology enabled genome-wide assessment of DNAm for many individuals at a moderate cost ^17,18^. Epigenome-wide association studies (EWAS) have shed light on many psychiatric disorders such as depression ^10^, anorexia nervosa, Alzheimer’s disease ^19,20^, or schizophrenia ^21^, complementing genetic research. Given their dynamic and modifiable nature, DNAm can be acquired or lost over the lifespan depending on environmental influences. Thus, epigenetic modifications may serve as biomarkers for gene x environment interactions, providing further insights into the molecular basis of OCD ^22^.

We recruited 384 participants from two German cities, Berlin and Bonn, to investigate the relationship between blood DNAm and OCD, which makes it the largest study to date. We first search for genomic loci showing differentially methylated sites between cases and healthy controls. We then computed a methylation profile score (MPS) to assess its classification power to differentiate cases from controls, as well as its association with treatment response and symptom severity (Y-BOCS scale).

## Results

### Epigenome-wide association study

To identify potential loci associated with OCD, we conducted a two-step case-control EWAS using samples recruited in Berlin for discovery and samples originating from Bonn for replication. This approach rendered in the Berlin sample a total of 188,488 DMPs with a nominal p-value < 0.05. These sites were moved forward to the replication stage using the Bonn samples. We identified 310 DMPs discriminating cases and controls with a corrected p-value for multiple testing q < 0.01 (Figure 1a).

**Figure 1:**
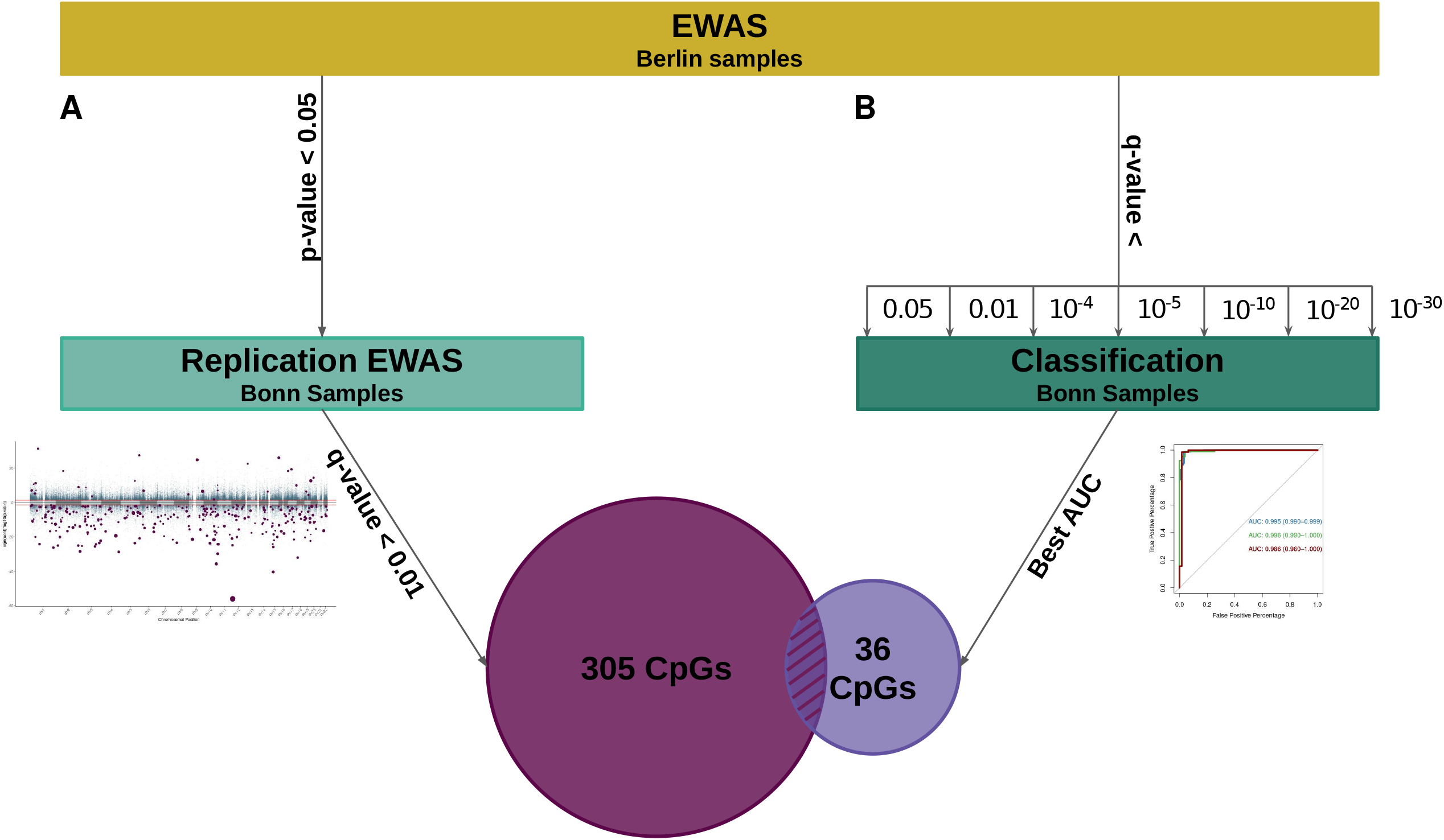
A schematic representation of our analysis. After the first EWAS only on the Berlin data set (discovery stage), probes are filtered based on two different approaches and replicated in the Bonn data set. Method A removed all probes with p-value > 0.05 in the discovery stage and run an EWAS on the remaining CpGs using the Bonn samples. The probes with q-value ≤ 0.01 (BH correction) were considered differentially methylated (DMP). Method B defined an a priori set of thresholds, q-values equals 0.05, 0.01, 10-4, 10-5, 10-10, 10-20, and 10-30, which were used to compute different MPS values. Next, the MPS was used as an independent variable to classify the Bonn samples and to select the best MPS based on the AU-ROC metric. The set of CpGs used to build the best MPS was selected. The intersection of both methods (12 CpGs) was then selected as the actual signals.

**Figure 2:**
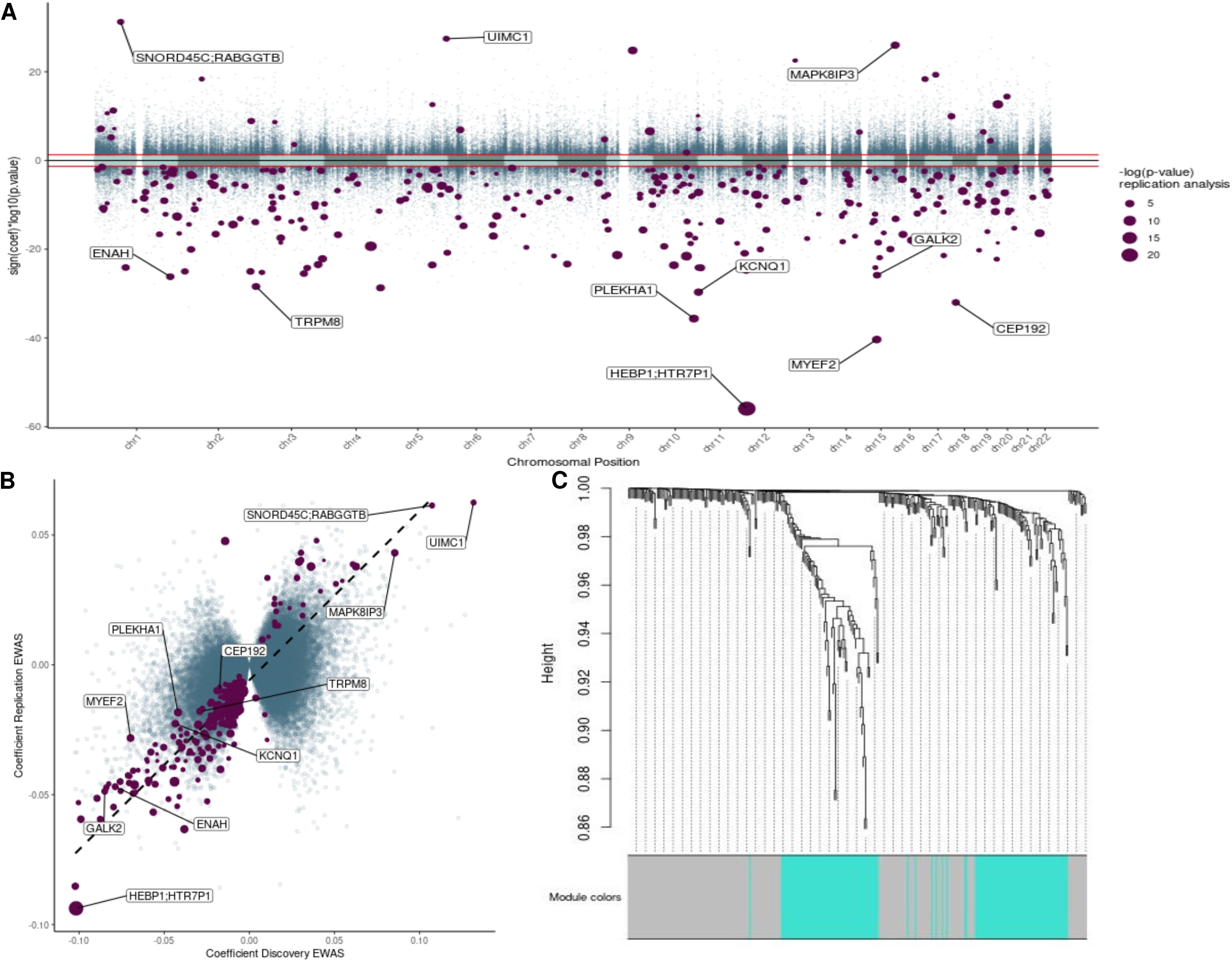
A) Miami Plot for the two-stage analysis. The X-axis is the genome position. Y-axis is the nominal p-value for the discovery EWAS on a logarithmic scale multiplied by the sign of the coefficient in the same analysis. Horizontal red lines define the threshold 0.05 of the discovery analysis. Purple dots are the 310 CpGs that were significant at the replication stage, the dot size is equivalent to the adjusted p-value in the replications stage on a logarithmic scale. B) Correlation of the discovery and replication stage. The X-axis shows the discovery coefficient, and Y-axis shows the replication coefficient. The purple dots represent the 310 CpGs that were significant at the end of the two-step approach; the dot size is equal to the adjusted p-value in the replication stage. The dashed line shows the trend of the linear model based on the purple dots. C) Cluster dendrogram. Branches refer to highly interconnected clusters of CpGs. Modules are represented by the colors in the horizontal bar.

We explored the correlation between the coefficients of the probes analyzed in the discovery and the replication stage (Figure 1b). This analysis showed that while the overall correlation for all 188,488 CpG sites was moderate (r = 0.42, p < 2.2×10^−16^), it was much stronger for the 310 DMPs in the replication stage (r=0.88, p < 2.2×10^−16^). Only five DMPs showed opposite effect directions between discovery and replication, therefore they were removed from further analysis (Supplementary table 1 and supplementary figure 1).

Of the 305 probes identified by our analysis, 241 were annotated to 233 genes based on the Illumina annotation. Gene Ontology (GO) analysis, using the R package missMethyl (49,50), for the same probes did not show any term enriched after multiple test corrections. Of note, five terms from the GO analysis showed a nominal p-value < 0.05 (Supplementary table 5).

### Network analysis identifies two different submodules

Given the complex nature and many pathways involved in OCD, we sought to search whether common patterns of methylation emerge among the 305 DMPs. Thus, we used WCNA which exploits correlations among probes and groups them into modules using network topology. After fitting several powers (β), we found that a power of ten approximated the best scale-free network for our co-methylation network (Supplementary figure 6). The adjacency matrix was then computed by using the optimal β and the methylation values. Based on the TOM dissimilarity measure, the hierarchical clustering yielded two consensus network modules, i.e. grey (n = 169, supplementary table 2) and turquoise (n = 136) (Figure 1c, supplementary table 2 and 3, and supplementary Figures 2 and 3).

Then we examined whether each module was associated with other phenotypes. To this end, we looked at the Pearson correlation coefficient and p-value of the association of the eigenvector of each module with OCD status, age, sex, city, smoking, and Y-BOCS. While both modules were highly correlated with OCD phenotype (turquoise: r = -0.88, p = 4x 10^−122^; grey: r = -0.79, p = 3 × 10^−78^), only the turquoise module was associated significantly with the Y-BOCS (r = -0.2, p = 2 × 10^−4^) (Supplementary Figure 6). Interestingly, the gray module better captured the differences in the origin of the samples.

### The methylation profile score offers predictive performance for sample classification

Considering that both submodules and the full set showed a strong correlation with OCD status, we attempted to derive a methylation profile score (MPS) by following a similar strategy to developing polygenic risk scores ^23^. To this end, we first constructed an MPS using only the 305 DMPs, which were confirmed in the replication stage (MPS_two-step_). We also computed an MPS for each module, i.e., turquoise (MPS_turquoise_) and grey (MPS_grey_).

The MPS_two-step_ was indeed statistically different between OCD patients and controls for both, the Berlin (p < 2.2×10^−16^) and the Bonn samples (p < 2.2×10^−16^), whereas the difference of MPS_two-step_ values between both cities for the control group (p = 0.269) and the OCD patients (p = 0.057) was not significant (Figure 3).

**Figure 3:**
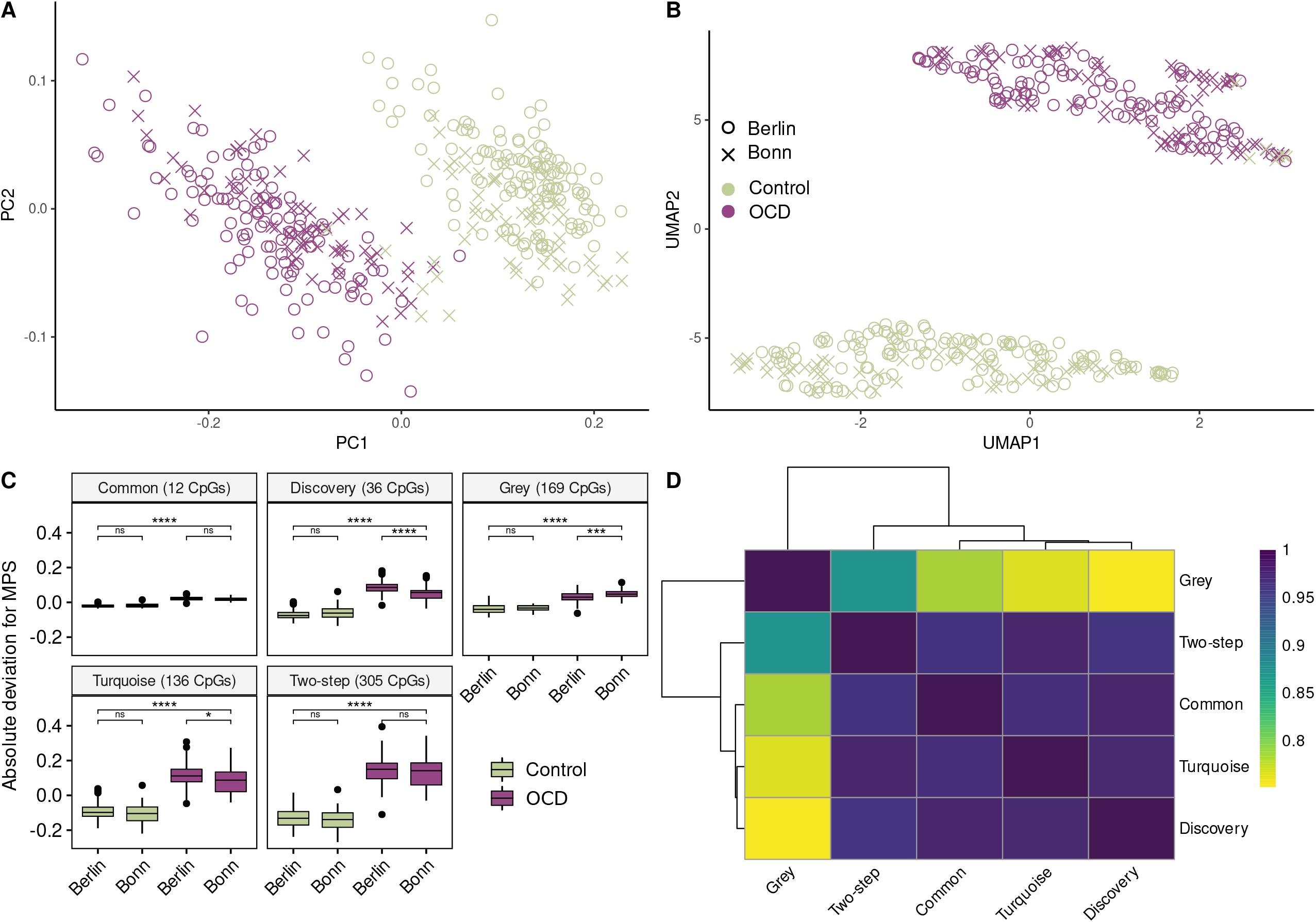
Projection of the samples into a two-dimensional space using A) PCA, and B) UMAP. The 12 CpGs found at the end of our analysis were used as input features. Purple data points are OCD patients and green are Controls. C) Each facet represents the deviation from the mean for each MPSs. The number of CpGs that were used to calculate the MPS is shown in parentheses. Horizontal brackets display the results of the t-test for the set. D) MPS correlation matrix. PC: Principal Component; UMAP: Uniform manifold approximation and projection; ns: not significant; *: p.value < 0.05; ** : p.value < 0.01; ***: p.value < 0.001; ****: p.value < 2×10^−16^.

The lack of an independent third validation cohort to test the MPS_two-step_ independently prompted us to consider an alternative strategy for constructing the MPS. Herein, we constructed several MPSs using DMPs based on an *a priori* set of 8 corrected p-value (q-values) thresholds (P_T_) obtained from the EWAS performed in the discovery stage (Berlin samples only). Finally, classification accuracy for each calculated MPS was examined in the Bonn data set, which did not contribute to this MPS and could be used to test out-of-sample classification accuracy. The best classification accuracy for the Bonn sample is achieved using probes with q-values < 1 × 10^− 20^ (AU-ROC_Berlin_ = 0.991, AU-ROC_Bonn_ = 0.968; Table 2). The MPS obtained for this threshold (MPS_discoverey_) contains 36 DMPs (Supplementary table 4 and supplementary figure 4), from which 12 are shared with the MPS_two-step_ and the MPS_turquoise_ (Table 2). For this reason, we also constructed an MPS containing only the common CpGs (MPS_common_) which also showed a good classification power (Figure 3 and table 2).

**Table 1:**
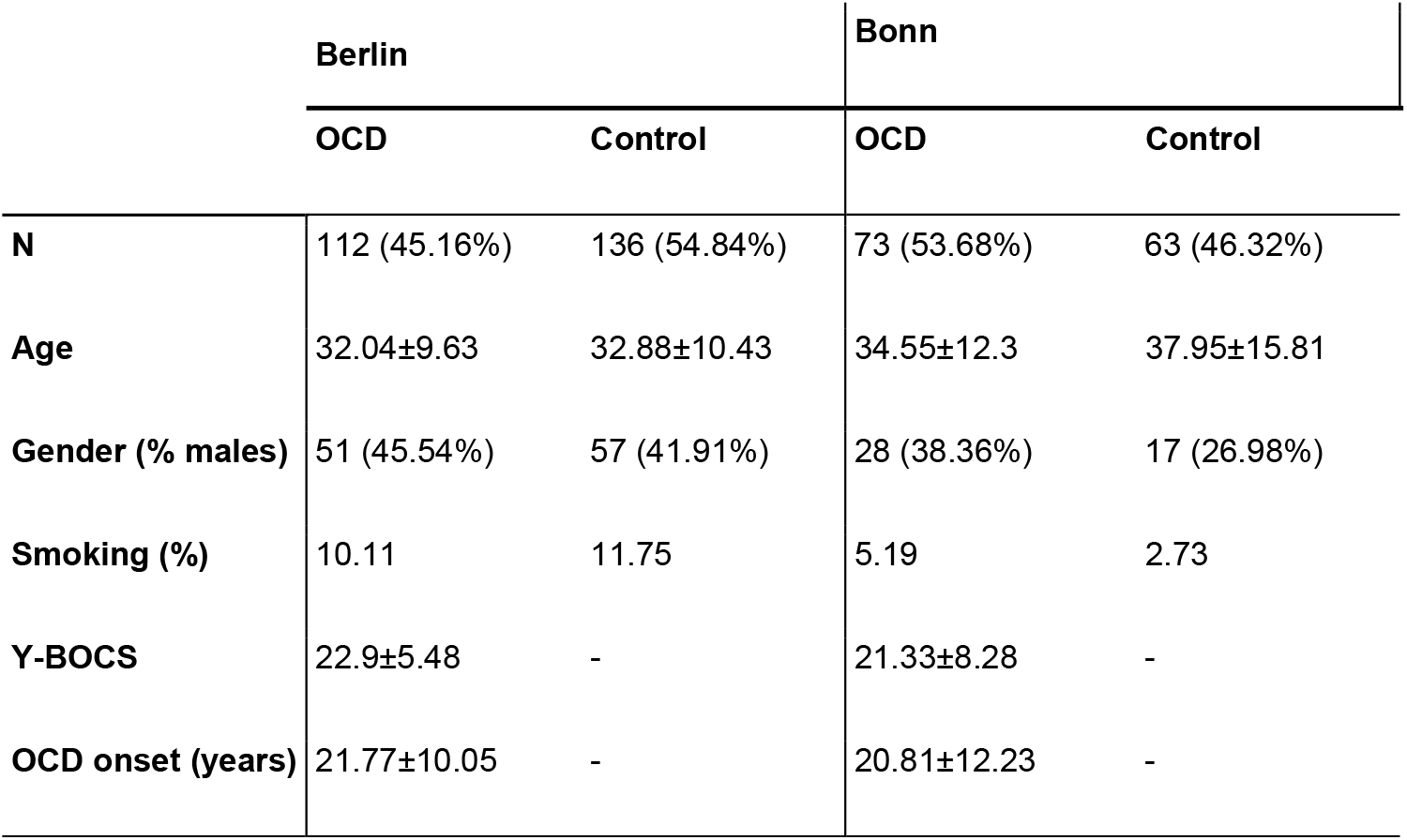
Cohort demographics. OCD: obsessive-compulsive disorder; Y-BOCS: Yale-Brown Obsessive-Compulsive Scale.

|  | Berlin |  | Bonn |  |
| --- | --- | --- | --- | --- |
|  | OCD | Control | OCD | Control |
| **N** | 112 (45.16%) | 136 (54.84%) | 73 (53.68%) | 63 (46.32%) |
| **Age** | 32.04±9.63 | 32.88±10.43 | 34.55±12.3 | 37.95±15.81 |
| **Gender (% males)** | 51 (45.54%) | 57 (41.91%) | 28 (38.36%) | 17 (26.98%) |
| **Smoking (%)** | 10.11 | 11.75 | 5.19 | 2.73 |
| **Y-BOCS** | 22.9±5.48 | - | 21.33±8.28 | - |
| **OCD onset (years)** | 21.77±10.05 | - | 20.81±12.23 | - |

**Table 2:** MPS properties. AU-ROC: Area Under the Receiver Operating Curve; Y-BOCS: Yale-Brown Obsessive-Compulsive scale; nCpGs: number of CpGs under the threshold selected; q-value: Bonferroni adjusted the p-value for the discovery study; r: Pearson correlation coefficient; p: p-value for the test.

|  |  | AU-ROC |  | Y-BOCS Correlation |  | nCpGs |
| --- | --- | --- | --- | --- | --- | --- |
|  |  | Berlin | Bonn | r | p |  |
| **q-values**<br>**threshold**<br>**for top-**<br>**down**<br>**Analysis** | 0.05 | 1 | 0.911 | - | - | 11,998 |
|  | 0.01 | 1 | 0.908 | - | - | 9,430 |
|  | 0.0001 | 1 | 0.909 | - | - | 4,902 |
| | $1 \times 10^{-5}$ | 1 | 0.915 | - | - | 3,568 |
| | $1 \times 10^{-10}$ | 1 | 0.88 | - | - | 808 |
|  | **$1 \times 10^{-20}$**<br>**(Discovery)** | 0.999 | 0.974 | 0.200 | 0.009 | 36 |
| | $1 \times 10^{-30}$ | 1 | 0.944 | - | - | 2 |
| **Two-step analysis** |  | 0.990 | 0.984 | 0.229 | 0.003 | 305 |
| **Grey** |  | 0.963 | 0.994 | 0.113 | 0.142 | 169 |
| **Turquoise** |  | 0.993 | 0.981 | 0.223 | 0.003 | 136 |
| **12 common CpGs** |  | 0.998 | 0.986 | 0.146 | 0.058 | 12 |

### Association between MPS, clinical variables, and treatment response

As indicated by Pearson correlation, the MPS_common_ was significantly associated with Y-BOCS scores across all OCD patients (r = 0.17, p = 0.023), indicating that a more severe symptom severity goes along with a higher epigenetic profile score. In the regression model assessing treatment response, we observed effects at the trend level for the Y-BOCS baseline score (β = - 3.108, t = -1.96, p = 0.053) and the Y-BOCS baseline by MPS_common_ interaction (β = -2.78, t = - 1.74, p = 0.086). To follow up on this interaction, we ran separate analyses for patients with high and low MPS_common_ (median split: n = 56 low-scorers, n = 44 high-scorers). In MPS_common_ high-scorers, we found a significant effect of Y-BOCS baseline (β = -0.44, t = -3.09, p = 0.004) and a trend level association of the MPS_common_ (β = -0.28, t = -1.86, p = 0.070) with treatment response, indicating that patients with a higher score might show a better treatment response independently of baseline symptom severity. In MPS_common_ low-scorers, there were significant effects of Y-BOCS baseline (β = -0.32, t = -2.43, p = 0.019) and medication (β = 0.35, t = 2.67, p = 0.010). Notably, we did not observe any significant effects of age or gender in all analyses (p > 0.05). Moreover, there was no significant association between MPS_common_ and Y-BOCS baseline score in the treatment subsample (r = 0.09, p = 0.37), potentially due to sample size reduction.

### Functional Annotation

We conducted a focused literature search on the 12 common CpGs identified in both MPS approaches, as they may represent true signals involved in the disease process operating in OCD as the GO analysis did not reveal clear supporting evidence for functional terms that may be relevant or previously associated with OCD. To this end, we first mapped each CpG to the closest gene and gene position (Table 3).

**Table 3:** Biological annotation and summary statistics for the 12 common CpGs. q-value: Bonferroni adjusted the p-value

|  | Position | Gene | q-value<br>discovery | Coefficient<br>discovery | q-value<br>replication | Coefficient<br>replication |
| --- | --- | --- | --- | --- | --- | --- |
| **cg17232014** | chr12:13153193 | HEBP1;<br>HTR7P1 | $7.38 \times 10^{-51}$ | -0.1 | $7.47 \times 10^{-24}$ | -0.09 |
| **cg01647172** | chr10:124146007 | PLEKHA1 | $1.45 \times 10^{-30}$ | -0.04 | $1.68 \times 10^{-6}$ | -0.02 |
| **cg13959110** | chr15:48466199 | MYEF2 | $2.78 \times 10^{-35}$ | -0.07 | $5.65 \times 10^{-6}$ | -0.03 |
| **cg00382572** | chr11:2574042 | KCNQ1 | $1.32 \times 10^{-24}$ | -0.04 | $1.08 \times 10^{-5}$ | -0.02 |
| **cg06215939** | chr16:1755402 | MAPK8IP3 | $6.45 \times 10^{-21}$ | 0.09 | $1.55 \times 10^{-5}$ | 0.04 |
| **cg20469575** | chr4:169122189 | | $1.26 \times 10^{-23}$ | -0.07 | $3.63 \times 10^{-5}$ | -0.05 |
| **cg19069918** | chr2:234921635 | TRPM8 | $2.53 \times 10^{-23}$ | -0.03 | $5.43 \times 10^{-5}$ | -0.02 |
| **cg25195309** | chr1:225766155 | ENAH | $4.12 \times 10^{-21}$ | -0.08 | $8.89 \times 10^{-5}$ | -0.05 |
| **cg07397958** | chr15:49476141 | GALK2 | $8.86 \times 10^{-21}$ | -0.08 | $9.63 \times 10^{-5}$ | -0.05 |
| **cg16449667** | chr18:13024185 | CEP192 | $6.44 \times 10^{-27}$ | -0.02 | $1.14 \times 10^{-4}$ | -0.01 |
| **cg21812670** | chr1:76251636 | SNORD45C;<br>RABGGTB | $3.66 \times 10^{-26}$ | 0.11 | $6.43 \times 10^{-4}$ | 0.06 |
| **cg19755108** | chr5:176434079 | UIMC1 | $2.30 \times 10^{-22}$ | 0.13 | $7.54 \times 10^{-4}$ | 0.06 |

The highest association was found for the CpG cg17232014, which shows a substantial hypomethylation in OCD patients compared to controls. This CpG maps to a transcription start site (TSS) for two genes: Heme Binding Protein 1 (*HEBP1*) and the 5-Hydroxytryptamine Receptor 7 Pseudogene 1 (*HTR7P1*), most commonly known as serotonin receptor pseudogene (Supplementary figure 7). Although the functional consequence of the decreased methylation at this TSS is not fully understood yet, it likely results in an elevated gene expression of either *HEBP1* or *HTR7P1* or both.

Next, we observed that some of the associated CpGs were located close to genes linked to glucose metabolism. Thus, the cg01647172 is mapped to the 5’ untranslated region of the gene Pleckstrin Homology Domain Containing A1 (*PLEKHA1*) and is found hypomethylated in OCD patients. Likewise, we observed that the hypomethylated CpG cg00382572 position is assigned to the KCNQ1 gene coding for the Kcnq1 potassium channel, which is located in the pancreas and has been also associated with diabetes ^24–29^. Finally, the cg19069918 is located near the gene *TRPM8*, which has been long studied as a cancer biomarker, particularly in pancreatic cancer ^30^.

The next set of CpGs was annotated to genes involved in different processes related to resident cells of the brain. Thus, the probe cg06215939 is found hypermethylated at the TSS of the Mitogen-Activated Protein Kinase 3 gene (*MAPKIP3*) predicting a reduction in gene expression. For cg21812670, the methylation was found to be increased in OCD patients. This position is located at the TSS of the gene coding for the Rab geranylgeranyl transferase (*RABGGTB*) which is essential for synaptic vesicle release ^31^. Along these lines, the cg13959110 is located in the gene coding for the brain myelin expression factor 2 (*MYEF2*). This gene is a transcriptional repressor of the myelin basic protein gene (*MBP*) that has been involved in myelin homeostasis. Another CpG, cg25195309 is located in the Enable Homolog gene (*ENAH*). The function of this gene has been linked to actin polymerization in neurons ^32^. Herein, neurons lacking these proteins cannot perform neuritogenesis in the developing cortex ^32^.

## Discussion

In the present study, our primary goal was to identify changes in DNAm associated with OCD status. Following a discovery and replication strategy, we identified 305 CpGs that were differentially methylated between cases and controls. Using these 305 DMPs, or a subset of them, allowed us to classify cases and controls accurately. Importantly, similarly, high classification accuracy was reached when we applied a different analytical strategy using the strongest disease-related DMPs signals of the Berlin sample to predict caseness in the independent sample from Bonn. Both analytical strategies converged on 12 common CpGs deserving further scrutiny. Finally, we found a significant association of a methylation score based on these common 12 CpGs with OCD symptom severity, as well as a trend level association with treatment response to CBT in OCD patients with high MPS, indicating that patients with larger values show better treatment response. This latter result might allow MPS to be used as a biomarker for predicting treatment response in OCD from a translational perspective.

While EWAS has already led to important advances in other neurological and psychiatric disorders, it is still early days for OCD epigenetics ^33,34^. For example, a study on the Chinese Han population reported 8,417 DMPs in the blood of 65 cases and 96 controls ^34^. In addition, the comparison of DNA methylation in the saliva of 59 patients with OCD and 54 controls of European origin identified nine genes with methylation changes related to OCD and ADHD which however did not survive multiple testing correction ^33^. In 2022, Shiele et al. reported nine genome-wide significant DMPs mapping to several microRNAs and pseudogenes in the saliva of 68 OCD patients and 68 controls of European origin ^35^. Importantly, we could not identify any overlapping signal in our datasets.

In this regard, a strength of our study is the two-step approach in which we treated Berlin and Bonn samples as independent cohorts. As a result, we were able to avoid the “winner’s course” in our analysis, i.e. overestimation of small effect sizes in underpowered cohorts. Although our sample size might seem underpowered, we defined the expected number of false positive signals that will arise from our study design following the methodology described by Jiang et al. ^36^. Thus, after permuting the samples to estimate the FP rates, on average, 32,711 probes would be significant after the discovery step, which is in agreement with the theoretically expected (0.05 × 632,997 ≈ 31650). In addition, the second step would not report any significant probe under the threshold imposed. Consequently, the overall FDR was 0.003%, which corresponds to approximately 21 false DMPs after the replication step. Therefore, we assume that genuine signals among the 305 CpGs identified in our study are included. Supporting this assumption, our analytical strategy converged on 12 common probes out of the 305 DMPs that may represent true pathophysiological processes involved in OCD.

Pathway search did not lead to the identification of obvious candidate pathways for OCD, but the disgenet ^37,38^ tool and literature search revealed that genes near the 12 CpGs have been linked to diseases like diabetes, Parkinson’s disease (PD), ADHD, and multiple sclerosis (MS). Interestingly, the pathogenic processes involving these genes are also linked to OCD, including glucose metabolism, the dopaminergic/serotonin system, and neuronal function. For glucose metabolism, we found that the PLEKHA1 locus has been associated with type 1 and type 2 diabetes mellitus (T1/2DM) and age-related macular degeneration (AMD) ^39,40^. In AMD, previous research has shown that TAPP1, a PLEKHA1 protein product, works as an activator of lymphocytes, indicating that PLEKHA1 plays a role in inflammation. Interestingly, increasing evidence has shown that inflammatory pathways are common pathogenetic mediators in the natural course of both types of diabetes that involve the activity of PLEKHA1 ^41^. For KCNQ1, research has shown that overexpression of the ion channel in mouse-derived pancreatic β-cells leads to an impairment in insulin secretion stimulated by glucose and pyruvate ^24^. Lastly, rats with deletion of the TRPM8 gene showed reduced insulin levels in serum due to enhanced insulin clearance in the liver. This was caused by afferent fibers innervating the hepatic portal vein, which is critical for metabolic homeostasis ^42^. Importantly, this latter mechanism also seems to be the intersection connecting the nervous system with the metabolism of glucose and insulin. Hence, our data suggest that an underlying dysregulation in insulin/glucose metabolism may drive, at least in part, the symptoms and the disease processes occurring in OCD patients. Unfortunately, we did not have serum samples from patients before and after therapy to analyze whether glucose and insulin homeostasis changed after treatment.

Besides insulin and glucose metabolism, we also identified several genes involved in brain function. For example, both genes near cg17232014 on chromosome 12 HEBP1 and HTR7P1 have been associated with brain phenotypes. Thus, increased expression of HEBP1 in the brain has been linked to neurotoxicity ^43^ and neuroinflammation ^44^. HTR7P1, although this is a pseudogene that does not translate into protein, genetic variants in HTR7P1 have been associated with neurological and growth phenotypes in children ^45^.

In our study, we identified several signals that support the sweet-compulsive brain hypothesis ^46^. This hypothesis states that abnormal dopaminergic transmission in the striatum may perturb insulin signaling sensitivity in OCD patients. Deep brain stimulation in patients with OCD supports the hypothesis that dopamine transmission affects glucose and insulin metabolism in the brain. Interestingly, non-diabetic OCD patients seem to have an increased hepatic and peripheral insulin sensitivity ^47^, supporting our findings on PLEKHA1, KCNQ1, and TRPM8. Further reinforcing our brain-related genes and their connection with glucose and insulin homeostasis, research on insulin receptor signaling in the central nervous system showed that insulin receptor signaling regulates the maintenance of synapses. In addition, insulin receptor signaling contributes to the processing of sensory information, as well as structural plasticity triggered by external experience _48_.

Thus, it is tempting to speculate that environmental factors and experiences in OCD patients might potentially lead to the disruption of brain circuits important for OCD through, at least in part, disruption of the insulin receptor signaling and the dopamine and serotonin system. Additionally, OCD patients have difficulty practicing healthy habits like sports and eating healthy, so the comorbidity of both diseases might be expected.

In supporting our findings, three of our twelve most significant DMPs were found in a recent study comparing people with GAD, or OCD, with healthy controls of Chinese Han origin ^49^. These probes map to RABGGTB, MPK8IP3, and ENAH genes. To our knowledge, this is the first time that two different studies on methylation in OCD replicated each other’s results using populations of different ethnic backgrounds. Of note, Guo et al. used a similar approach and methodologies to analyze their data as in our study. Herein, DNAm is highly sensitive to batch effect and other factors that might increase the variability. Therefore, it is crucial to account for confounding factors when analyzing this kind of data set.

The correlation between MPS and OCD symptom severity highlights the potential clinical utility of epigenetic measures. Future studies should examine whether changes in symptom severity also go along with epigenetic modifications. Interestingly, we observed a trend-level association between MPS and treatment response in OCD patients, indicating that patients with the highest MPS showed better treatment response independent of baseline symptom severity. Among MPS-low scorers, there was no association with treatment response. Although we interpret this preliminary finding with caution, it may show that patients with high MPS exhibit features that make them benefit more from CBT than others, e.g., a larger environmental component contributing to their OCD.

Our results should be interpreted considering some important limitations. First, DNA extraction was done in Bonn for all samples including those derived from the Berlin sample. Consequently, Berlin blood samples were transported uncooled before DNA extraction, which may contribute to variation in the methylation analysis. However, our study considered this source of bias including the fact that we initially analyzed both samples independently. To avoid this source of technical bias, future studies should include cool transport of blood samples to the processing center or proceed locally with the DNA extraction before frozen transport to the analyzing center.

In summary, we identified 12 epigenome-wide significant CpGs for OCD using a robust statistical analysis of two German samples. The clinical validity of these CpGs is supported by the significant associations of our methylation profile score with OCD diagnosis, symptoms severity, and – at trend level – treatment response to CBT. Furthermore, genetic annotation contemplates a strong interaction of insulin and the dopaminergic system with OCD. Our findings thus support the role of epigenetic mechanisms in OCD and may help pave the way for biologically-informed individualized treatment options.

## Methods

### Patients and Controls

Biological samples were obtained from 185 patients with OCD and 199 healthy individuals who participated in the Endophenotypes of OCD (EPOC) study ^50,51^. The two recruitment centers, the Department of Psychology of Humboldt-University in Berlin and the Department of Psychiatry and Psychotherapy of the University Hospital in Bonn, enrolled and evaluated all participants according to the same protocols (Table 1). Healthy individuals from the general population were recruited through public advertisements. All participants came from European ancestry. Before recruitment, written informed consent was given by all participants, and monetary compensation was paid for their time. The study was performed following the revised Declaration of Helsinki and approved by the local ethics committees of Humboldt University and the University Hospital Bonn.

### Clinical evaluation

All participants were examined by trained psychologists using the Structured Clinical Interview for DSM-IV (SCID-I) ^52^. The severity of OCD symptoms was evaluated using the German version of the Yale-Brown Obsessive-Compulsive Scale (Y-BOCS) ^33,53^. Patients with OCD were included if they: (a) were free of any psychotic, bipolar, or substance-related disorder in the past or present (b) had not been treated with any neuroleptic drug during the past 4 weeks, and © had not used benzodiazepines 2 weeks before the study examination. Moreover, healthy participants were excluded if they (a) had taken any psychoactive drug in the past 3 months, (b) reported any Axis I disorder, or (c) had a relative with OCD.

Current or previous treatments were assessed in the patients’ group, in which approximately 50% had received pharmacotherapy, predominantly with SSRI. 79 OCD patients reported treatment with psychotropic medication in the past four weeks. A total of 25 patients had their treatments discontinued several weeks before baseline, and did not take any specific medications at the time of assessment. Another 98 patients were medication-naive, reporting no priory psychotropic medication. Four patients did not provide a medication status report. The majority of patients had one or more comorbid Axis I disorder, with major depressive disorder being the most common comorbidity (n = 41).

### Treatment subsample

A subsample of OCD patients completed individual CBT at a university outpatient unit at the Berlin study site (Hochschulambulanz für Psychotherapie und Psychodiagnostik der Humboldt-University). The CBT sessions were administered by licensed psychotherapists and conformed to the general conditions for psychotherapy in the public German health care system, typically consisting of 25 or more individual 50-minute sessions per week. Details about the treatment can be found in Bey et al. (2021) ^54^ and Kathmann et al. (2022) ^55^. For *n* = 100 patients (n = 54 female, n = 46 male), Y-BOCS data was available at pre- and post-treatment ^56^.

### Methylation arrays

Blood aliquots were obtained from all participants. Genomic DNA was isolated from whole blood and DNA concentration and purity were determined using the NanoDrop ND1000 spectrophotometer (Thermo Fisher Scientific, Waltham, MA, USA). All samples were of sufficient quantity and quality. 500 ng genomic DNA was used as input for the bisulfite conversion reaction using the EZ-96 DNAm Methylation-Lightning MagPrep Kit (Zymo Research Europe GmbH, Freiburg, Germany) with an elution volume of 15μl. Bisulfite-treated DNA was vacuum concentrated and resuspended in 10μl. A total of 4μl of the resuspension was used as input for the Infinium Methylation EPIC BeadChip (Illumina Inc, San Diego, CA, USA). All analysis steps were performed following the manufacturer’s instructions. The Illumina iScan was used for imaging the array and data were exported in .idat format.

### Data acquisition and Quality Control

The R (Bioconductor) Meffil ^57^ package was used throughout our pipeline to analyze the complete data set. All raw idat files were pooled together to run the quality control (QC) and normalization steps. Samples were removed if there was a mismatch between the estimated methylation sex and the gender provided by the participant, deviations from the mean value for control probes, or the median intensity for the methylated or unmethylated signal deviated more than three standard deviations (s.d.).

Probes were removed for further analysis if they mapped to a sex-chromosome, had a detection p-value below 0.05, beadcount lower than three, or were aligned to multiple locations in the genome according to Nordlund et al. ^58^. In addition, we removed the 10% of probes with the lowest variability to reduce the number of probes and multiple tests ^59^. In the end, 366 samples (189 controls and 177 cases) and 632,997 probes passed all our quality control filters and were used to normalize the methylation intensities.

Functional normalization ^57,60^ was applied to remove technical variation using 15 principal components (PCs) and an assessment center (Berlin/Bonn) as a fixed effect. Blood cell proportion was imputed using functionalities from meffil for each individual and used in the linear models to correct the methylation effect.

### Two-step EWAS

To analyze our data set, the two cohorts were initially kept separated (Berlin and Bonn). While the larger cohort from Berlin served as a discovery cohort in the EWAS, the Bonn cohort was used for replication.

Meffil uses the Independent Surrogate Variable Analysis (ISVA) method which allows for estimating confounding factors (CF) in methylation studies ^57,61,62^. Briefly, the ISVA uses the independent component analysis (ICA) method to model CFs as statistically independent variables in each probe analysis ^62^. Thus, ISVA provides a non-supervised framework for accounting for any CF.

Methylation status was compared between controls and OCD cases using a linear regression model. Adjustments were made for age ^63^, sex ^64^, smoking ^65^, cell composition ^66^, and surrogate variables calculated by meffil.

The current strategy for selecting CpGs for further analysis aims first to remove the maximum number of probes in the discovery step optimizing the minimum number of false negatives (p-value < 0.05). The replication step follows with a more restrictive adjusted p-value (q-value) threshold to select CpGs that are truly associated with the phenotype (Holm-Bonferroni q-value < 0.01). A similar strategy has been applied to genetics ^67^ and methylation ^10,20^ studies.

We estimated the false discovery rate (FDR) for our approach following the method suggested by *Jiang et a*l. ^36^:

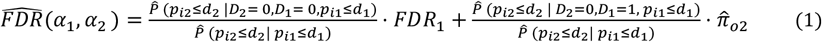

Briefly, a probe i with p_i1_ ≤ c_1_ will pass to the second stage, where p_i1_ is the p-value in the first stage and c_1_ is the threshold for the first stage. Following similar arguments for the second stage, p_i2_ ≤ c_2_, then we say that this probe has a significant difference in methylation values between cases and controls. At stage j, d_j_ is the smallest p-value for the probes that p_ij_ > c_j_, and the D_j_ is a binary variable that indicates whether there are actual differences between the cases and controls; D_j_ = 0 for no differences, and Dj = 1 for actual differences. The probability that a probe is significant after our two-stage approach when there are no real differences,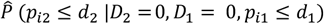, was estimated by permutating for 100 times the samples. The proportion of the true null hypothesis 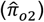 was estimated following the Storey method ^68^ and 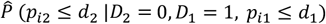 equals d_2_. Finally, 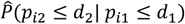 is the proportion of significant probes in the second stage. Our calculation for our setup yielded an FDR of 3.26×10^−5^.

### Weighted Correlation Network Analysis

Weighted correlation Network Analysis (WCNA) uses the pairwise correlation between variables to define clusters within the variables and to associate these clusters with other phenotypes.

The R package WGCNA was used for this purpose ^69,70^. Once the network was constructed, module detection was achieved by unsupervised clustering. WGCNA uses the dynamic tree-cut method to select the number of clusters given the hierarchical clustering for the adjacency matrix.

### Case-Control classification based on Methylation Profile Score

The methylation profile score (MPS) is a numerical value computed for each individual using a set of DMPs. Like polygenic risk scores in genetic studies ^71^, MPS improves classification capacity by leveraging methylation information on DNAm differences between cases and controls. The MPS for an individual i can be computed as

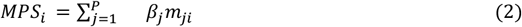

Where P is the number of CpGs, β_j_ is the coefficient for the association of the probe j to the phenotype, and m_j,i_ is the methylation status of probe j. Herein, a set of q-values thresholds was used to select the number of probes: 0.05, 0.01, 1×10^−4^, 1×10^−5^, 1×10^−10^, 1×10^−20^, 1×10^−30^, and 1×10^− 40^. For each threshold, an MPS was computed using the selected CpGse and then its classification capacity was tested using the Bonn sample as an independent dataset. AU-ROC for each threshold was computed using the R-package p-ROC.

### Clinical correlates and treatment analysis

To assess whether the OCD-related methylation profile is associated with symptom severity, we correlated our most reliable MPS (i.e. MPS_common_, see Results) with the Y-BOCS scores of all patients. In the treatment subsample, we also examined whether the MPS_common_ predicts treatment response by performing linear regression analysis with Y-BOCS baseline score, and the Y-BOCS baseline score × MPS_common_ interaction as independent variables, and pre-to-post change in Y-BOCS score as the dependent variable. Age, gender, and medication were included as covariates.

### Dimensionality reduction

A linear transformation algorithm and a non-linear transformation algorithm were used to reduce dimensionality. Principal component analysis (PCA) is the most popular linear transformation for dimensionality reduction. PCA estimates new coordinates that preserve the maximum variance of the dataset and projects the data points into the new orthogonal coordinate system. The base function prcomp in R was used to estimate the new PCs and projections. On the other hand, uniform manifold approximation and projection (UMAP) has become one of the most popular non-linear transformation algorithms. By using a framework that combines geometry and algebraic topology, UMAP can project a data set into two dimensions and reflect distances between points. The function umap in R was used to obtain the new coordinates.

## Data Availability

All data produced in the present study are available upon reasonable request to the authors

## Acknowledgments

This work was supported by the German Research Foundation (DFG) grants numbers: [KA815/6-1] to Norbert Kathmann, [WA731/10-1], [WA731/15-1] to Michael Wagner, and [RA1971/8-1], [RA1971/7-1] to Alfredo Ramirez.

## Author contributions

Drafting of the manuscript: RCM, AR, KB, AP, NK, and MW; epigenetic and phenotype data acquisition: BE, BR, and JK.; bioinformatics analysis: RCM. Treatment analysis: KB, and NK.; Study design: AR, MW.; Obtaining funding: AR, MW, NK, and, AP Critical revision of the manuscript: all authors.

