## Supplementary Figures for "Epigenome-wide analysis identifies methylome profiles linked to obsessive-compulsive disorder, disease severity, and treatment response"

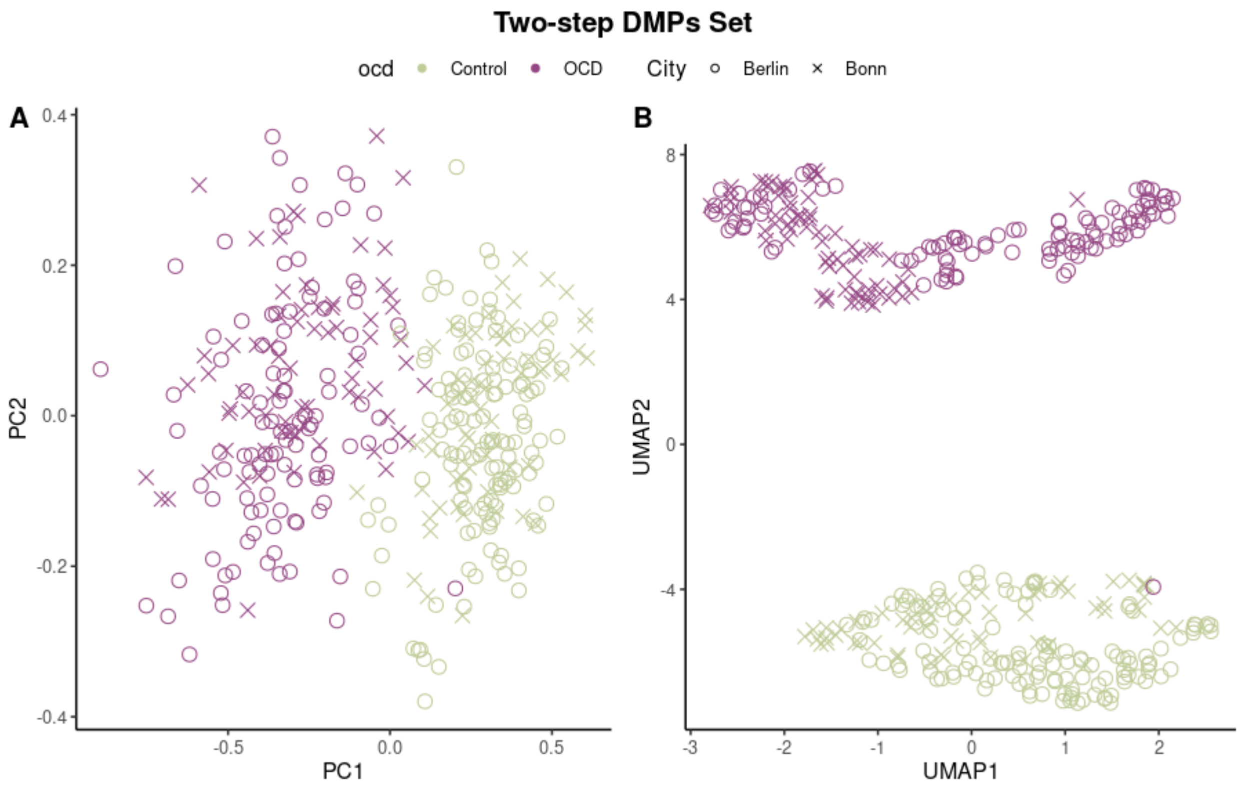


Figure 1. Dimensionality reduction using PCA and UMAP algorithm for the samples using the two-step (305) CpGs.

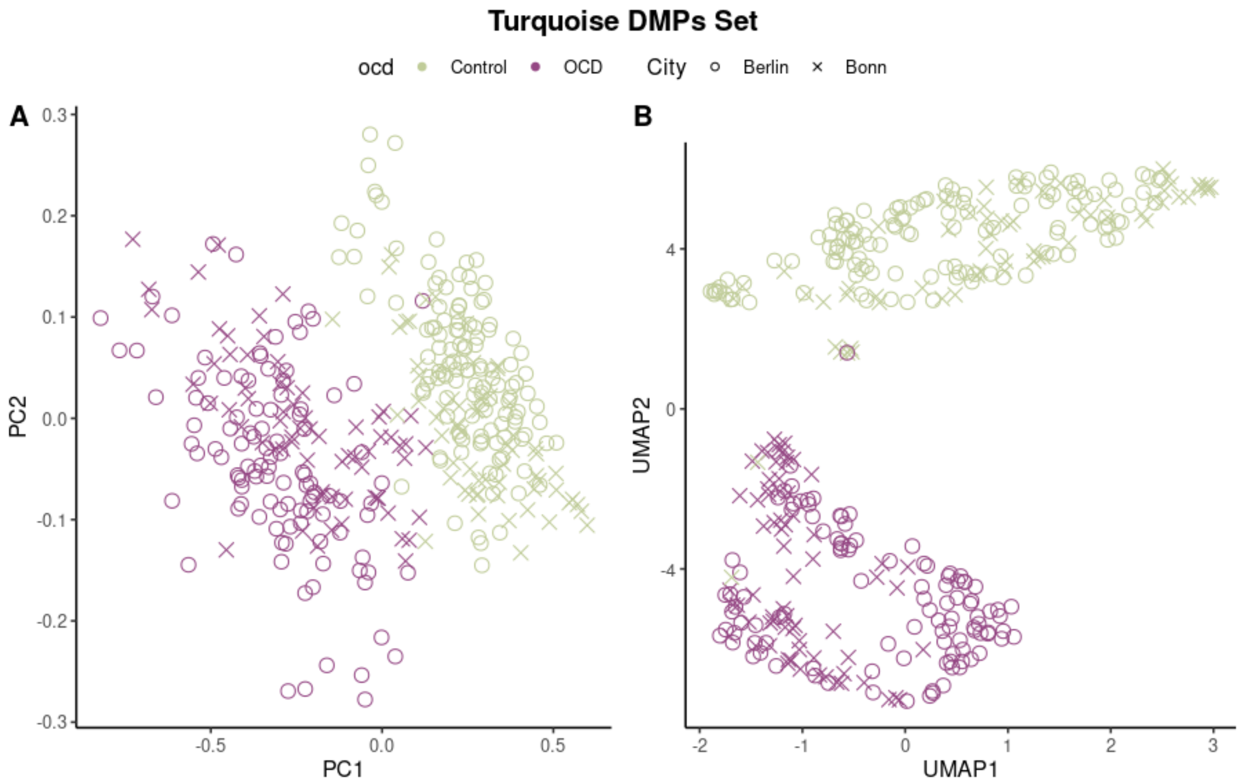


Figure 2. Dimensionality reduction using PCA and UMAP algorithm for the samples using the turquoise (136) CpGs.

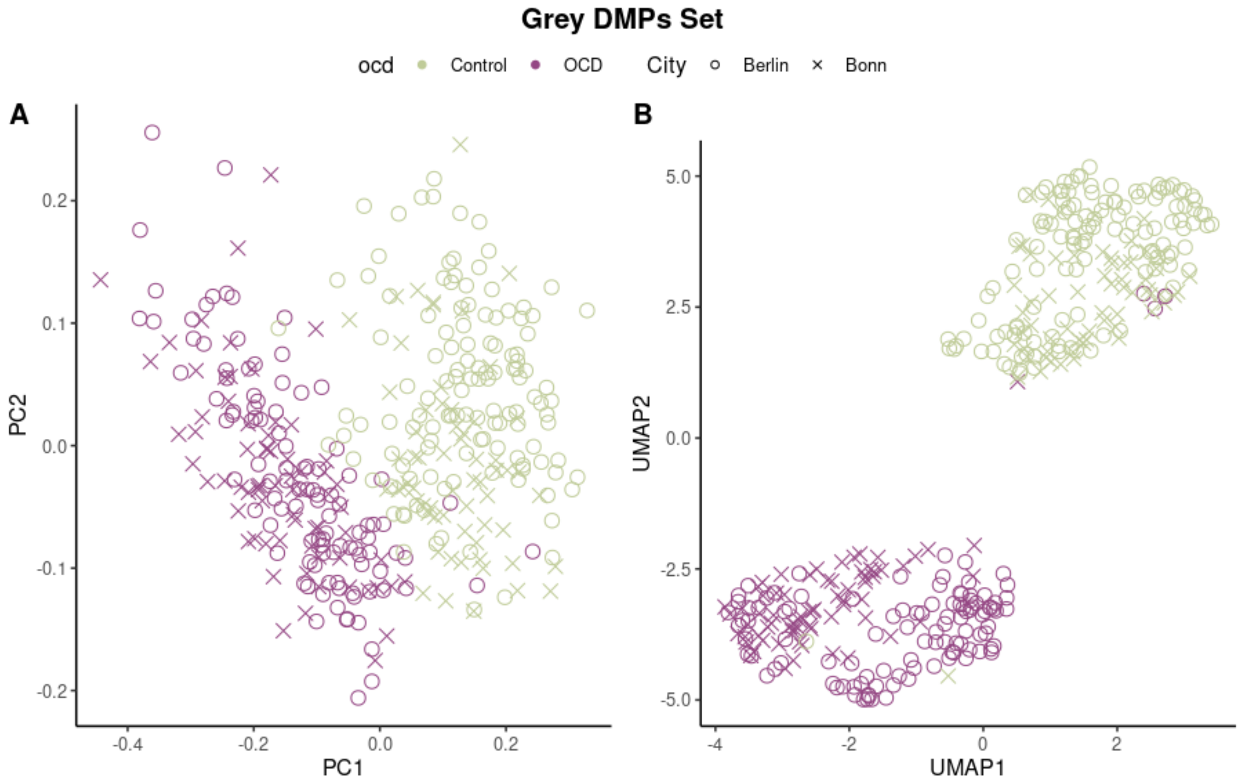

Figure 3. Dimensionality reduction using PCA and UMAP algorithm for the samples using the grey (169) CpGs.

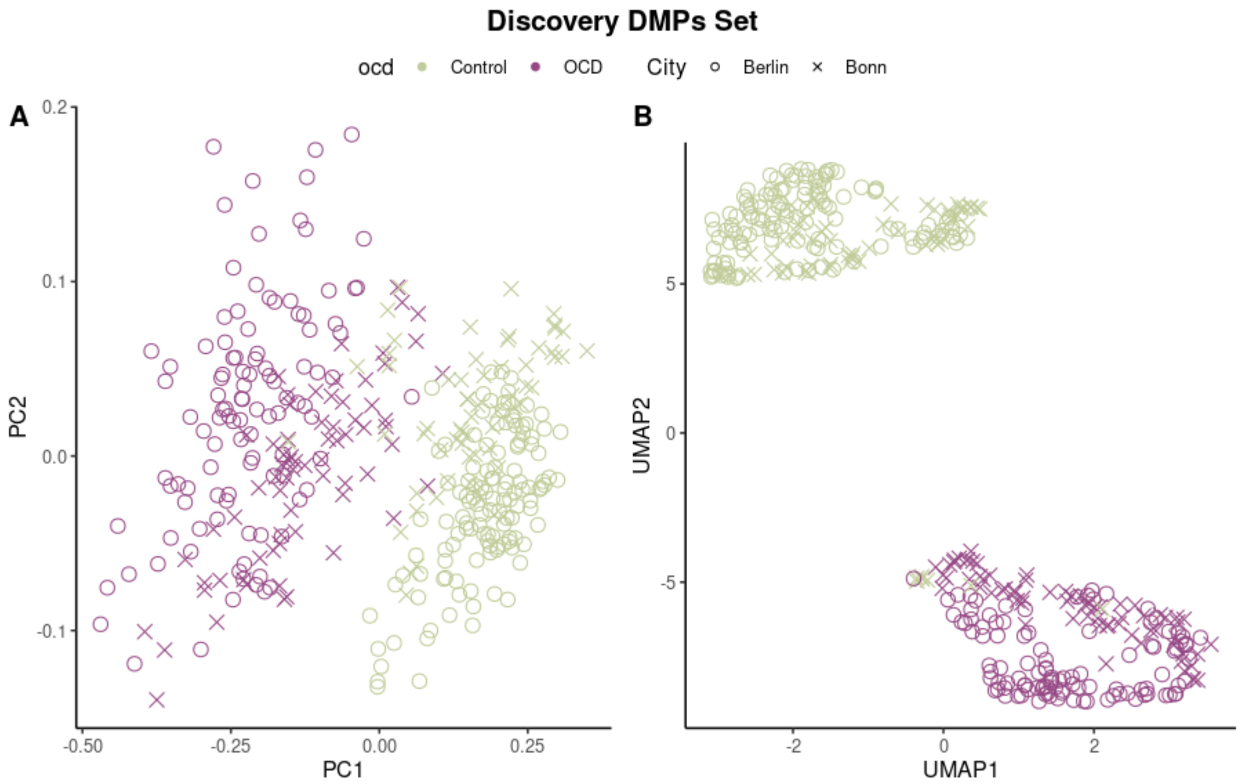


Figure 4. Dimensionality reduction using PCA and UMAP algorithm for the samples using the discovery (36) CpGs.

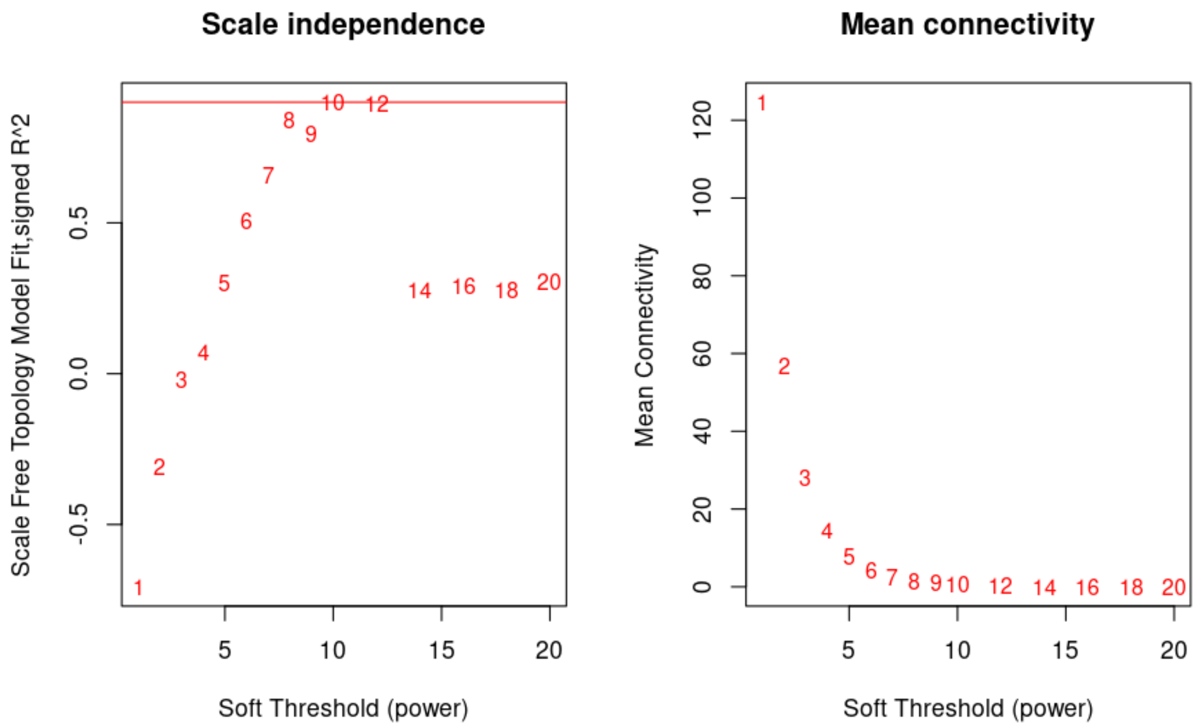
Scale independence
Scale Free Topology Model Fit,signed R^2
Soft Threshold (power)
Mean connectivity
Mean Connectivity
Soft Threshold (power)

Figure 5.

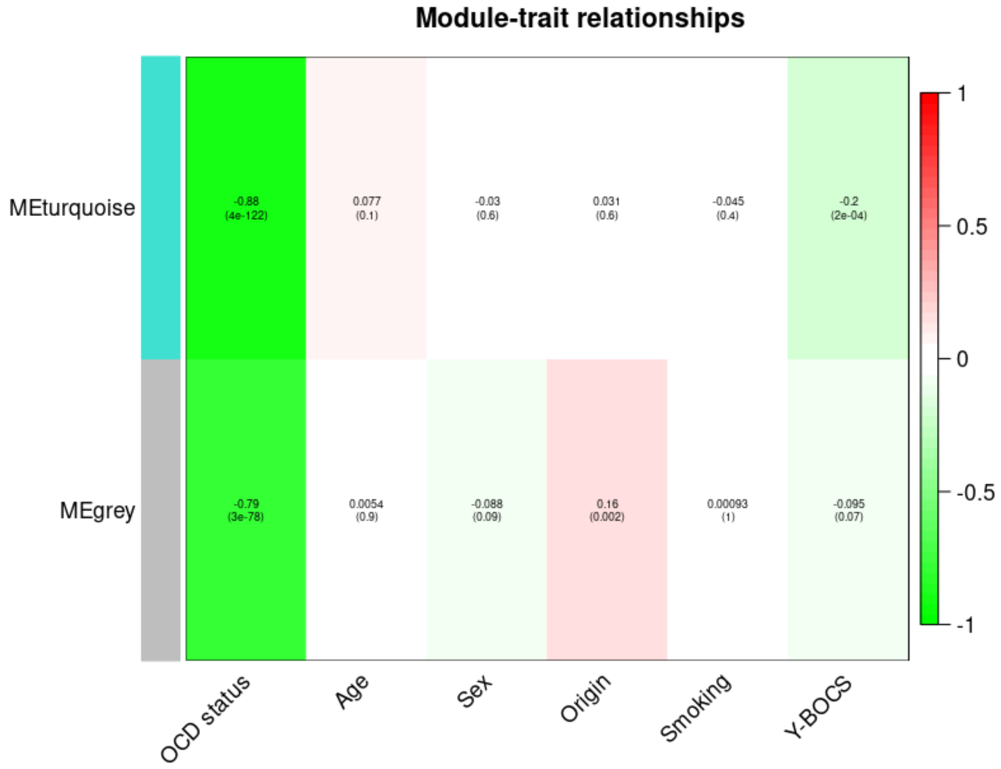
Module-trait relationships
MEturquoise
MEgrey
-0.88 (4e-122)
0.077 (0.1)
-0.03 (0.6)
0.031 (0.6)
-0.045 (0.4)
-0.2 (2e-04)
-0.79 (3e-78)
0.0054 (0.9)
-0.088 (0.09)
0.16 (0.002)
0.00093 (1)
-0.095 (0.07)
OCD status
Age
Sex
Origin
Smoking
Y-BOCS

Figure 6: Correlation of each submodule in the WCNA with different variables.

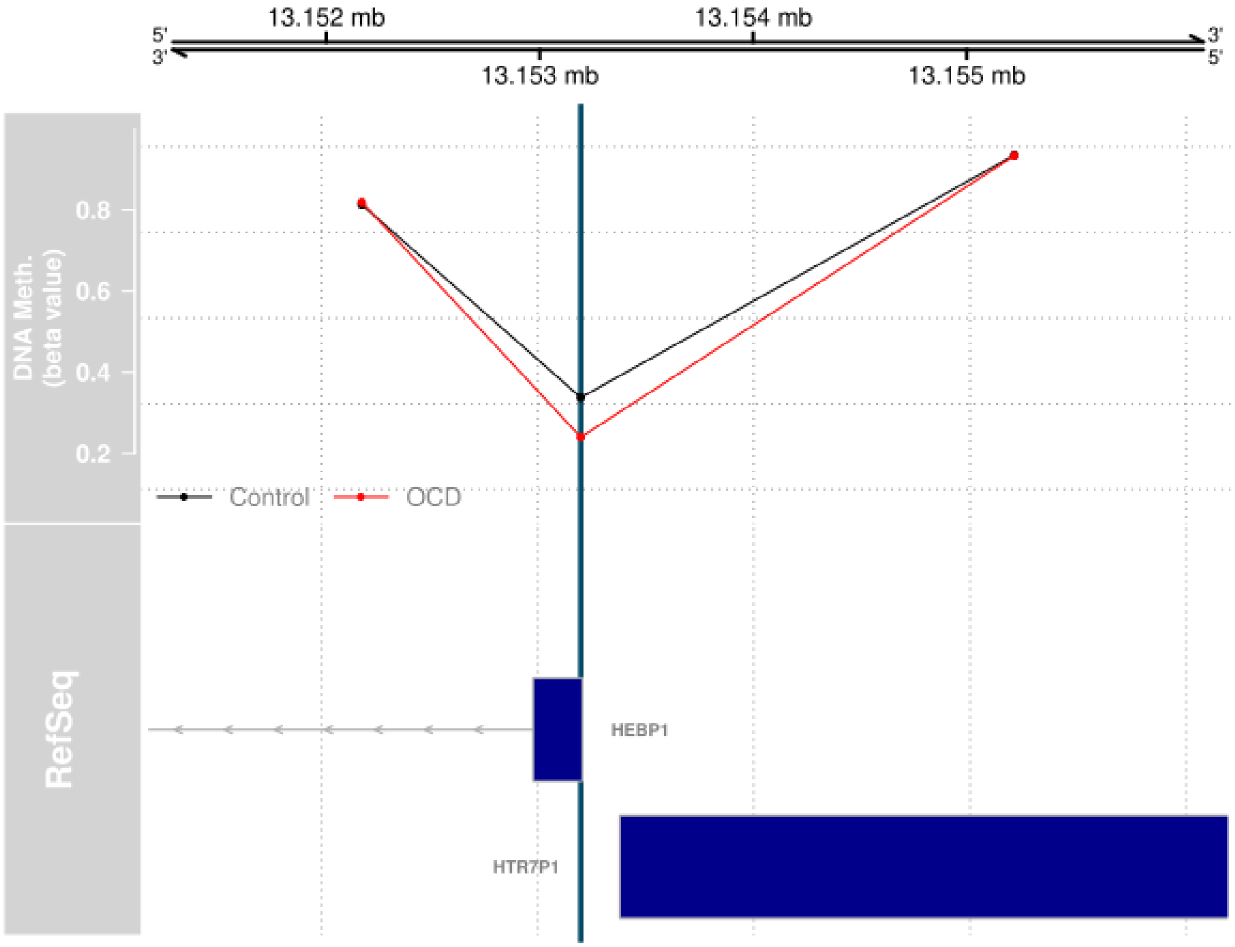


Figure 7: 5K locus of chromosome 12 spanning from 13.151Mb to 13.156Mb. The first panel shows the average methylation for the OCD samples (red dots) and controls (black dots). The cg17232014 probe (green vertical line) lies in a Transcription Start Site for both genes HEBP1 and HTR7P1. This probe is highly differentially methylated in both studies, the discovery and the replication study.

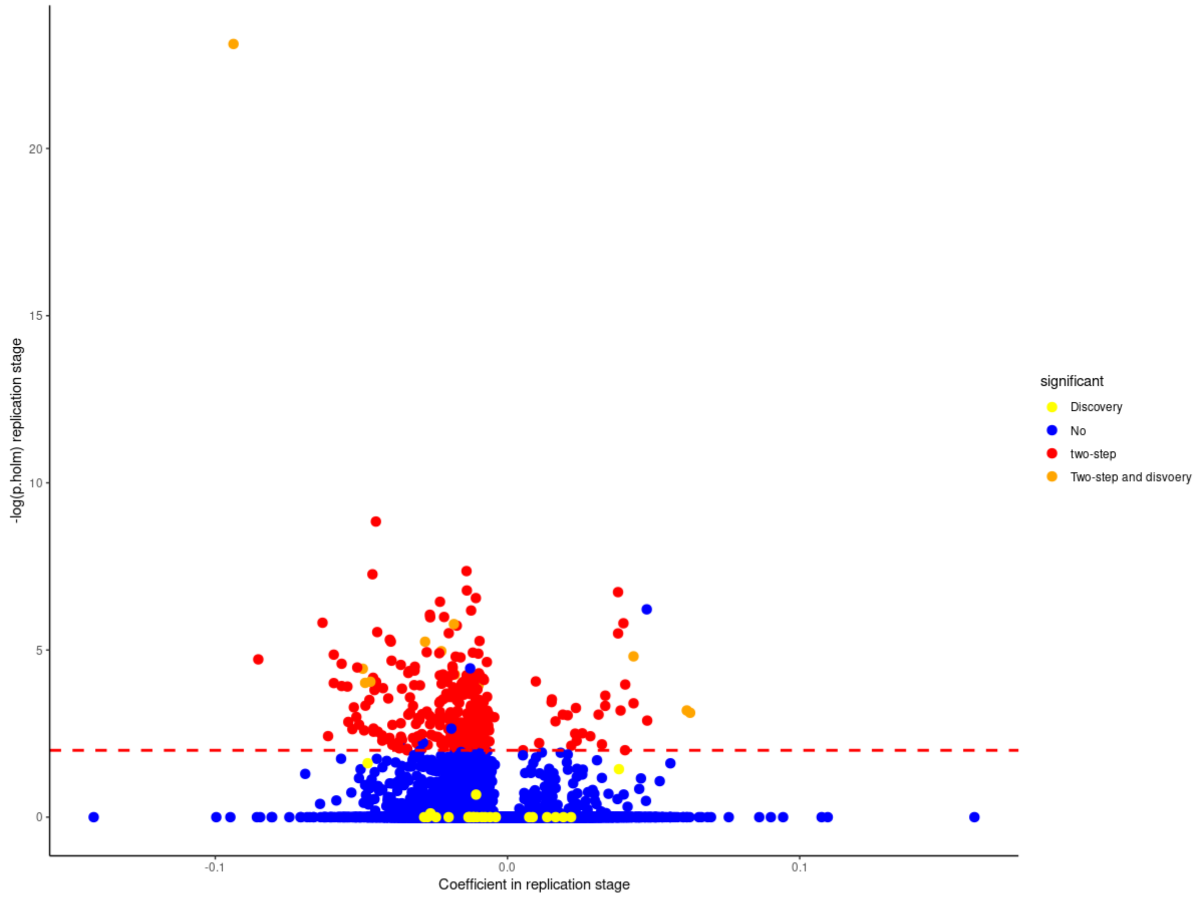


Figure 7: Volcano plot for the replication EWAS. Yellow dots were found in our discovery set but not in the replication step (24 probes). Blues dots are not significant in either of the analysis. Red dots are the DMPs from our two-step analysis but were not found in the discovery (293 probes). And the orange probes are the common CpGs (12 probes).
